# Prolonged SARS-CoV-2 replication in an immunocompromised patient

**DOI:** 10.1101/2020.09.20.20196899

**Authors:** Ji Hoon Baang, Christopher Smith, Carmen Mirabelli, Andrew L. Valesano, David M. Manthei, Michael Bachman, Christiane E. Wobus, Michael Adams, Laraine Washer, Emily T. Martin, Adam S. Lauring

## Abstract

We describe a case of chronic COVID-19 in a patient with lymphoma and associated B-cell immunodeficiency. Viral cultures and sequence analysis demonstrate ongoing replication of infectious SARS-CoV-2 virus for at least 119 days. The patient had three admissions related to COVID-19 over a four-month period and was treated twice with remdesivir and convalescent plasma with resolution of symptoms. The patient’s lack of seroconversion and prolonged course illustrate the importance of humoral immunity in resolving SARS-CoV-2 infection. This case highlights challenges in managing immunocompromised hosts, who may act as persistent shedders and sources of transmission.

## Introduction

Since the emergence of SARS-CoV-2, the causative agent of COVID-19, there has been an ongoing effort to define viral kinetics, host immune responses, and transmission dynamics. In individuals who develop symptoms, the average incubation period is approximately 5 days (IQR 2-7). The viral load in the respiratory tract peaks around the time of symptom onset and shedding of infectious virus occurs for 2-3 more days ^1^. While peak viral loads in asymptomatic and symptomatic individuals are similar, those with symptoms appear to shed viral RNA in greater quantities for longer periods. Individuals can test positive for nucleic acid for up to 6 weeks after symptom onset ^2^. When evaluated, infectious virus is generally not detected after greater than 7 days after symptom onset and contact tracing studies suggest that individuals are most infectious within 5 days of symptoms ^3,4^. The majority of infected individuals experience an illness lasting 1-2 weeks. For those requiring hospitalization, the median time from symptom onset to admission is 7 days (IQR 3-9) ^1^. The resolution of symptoms often coincides with seroconversion, with IgG levels increasing between 7-14 days post-infection ^5^.

Immunocompromised individuals are under-represented in most of these studies, and patients with primary or secondary immunodeficiencies may differ in their degree of shedding, kinetics of immune clearance, and disease severity. There is also likely to be considerable variability based on the type and severity of underlying immune deficit ^6^. Here, we describe the clinical and virological course of a patient with functional B-cell immunodeficiency and COVID-19.

### Case

A 60-70 year old man with a history of refractory mantle cell lymphoma (immunohistochemistry positive for CD20, CD5, BCL2, cyclin D1 and SOX11; lambda-restricted, CD5-positive B-cell population by flow cytometry) presented to the emergency department with a one week history of epistaxis and a cough productive of blood-streaked sputum. Chemotherapy for his lymphoma was ongoing as part of a clinical trial and included mosunetuzumab (a CD20-CD3 T-cell engaging bispecific antibody) in combination with cyclophosphamide, doxorubicin, prednisone and polatuzumab vedotin (a CD79b-directed antibody-drug conjugate). He was afebrile and did not require supplemental oxygen. His chest radiograph was without abnormality. A nasopharyngeal (NP) swab was positive for SARS-CoV-2 by reverse transcription polymerase chain reaction (RT-PCR, Diasorin assay).

He was admitted to the hospital (day 7 of illness) for monitoring in the setting of chemotherapy-associated neutropenia and severe thrombocytopenia. He received a platelet transfusion and his presenting symptoms improved. With resolution of his cytopenias, he was discharged to his home 6 days later (day 13, Figure 1A).

**Figure 1:**
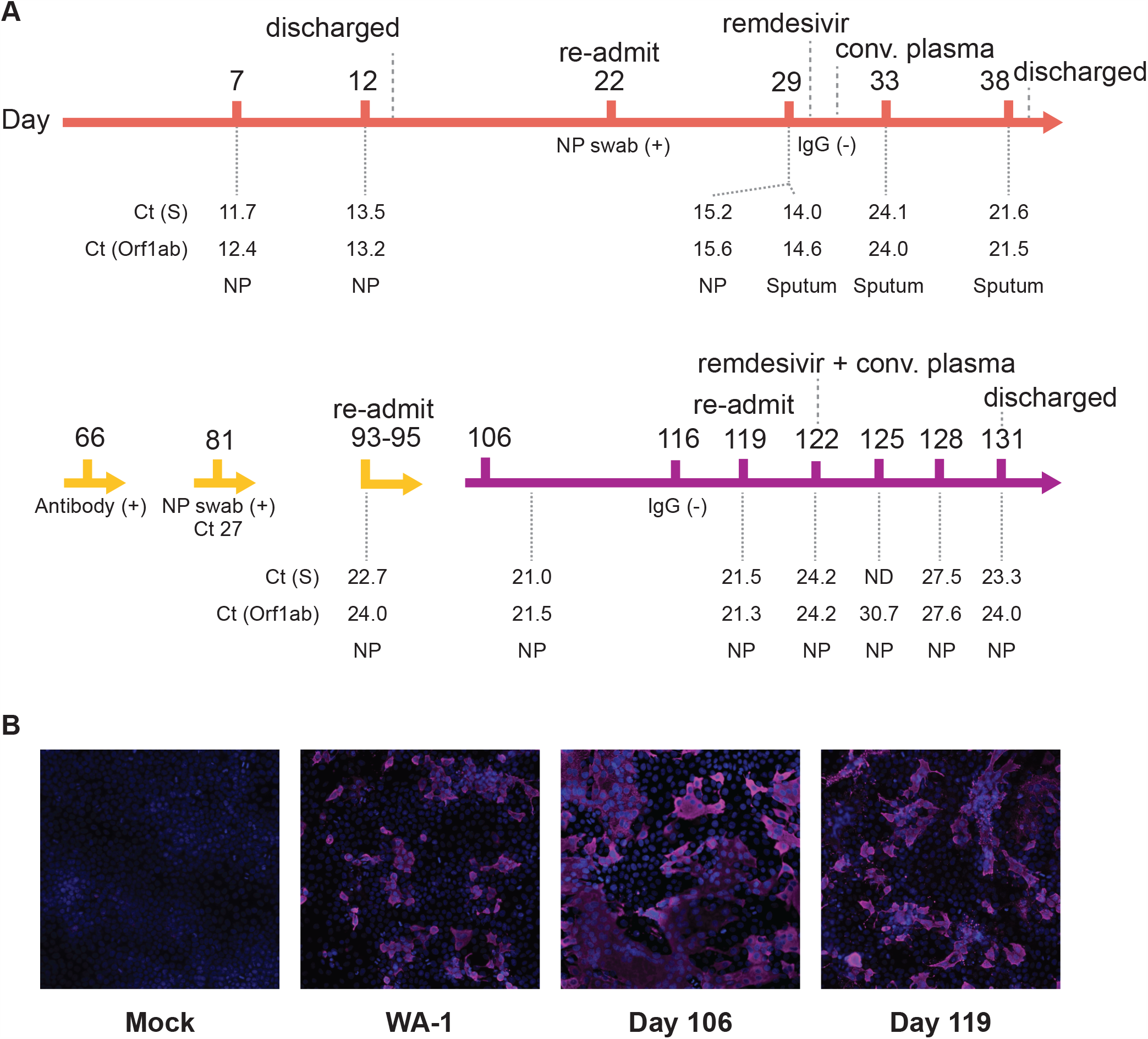
(A) Timeline of case. Color is by stage of infection, orange (initial presentation); yellow (largely outpatient); purple (recrudescence and readmission). Key events and treatments are shown above the timeline and key laboratory values are shown below it. ND = not detected, NP = nasopharyngeal swab, Ct = cycle threshold, S = spike, Orf1ab = open reading frame 1a and 1b. (B) Immunofluorescence microscopy of viral cultures. The patient’s samples (days 106 and 119) and a positive control (WA-1 strain) viral stock were cultured on Vero E6 cells until cytopathic effect was observed. Huh-7 cells were infected with cell-free supernatants from the Vero E6 cultures and stained 48 hours later with Hoechst (blue, nuclei) and an anti-SARS-CoV-2 nucleocapsid antibody (magenta).

Over the following week, he continued to have fatigue and a mild cough, but remained afebrile. He began to experience mild shortness of breath and returned to the emergency department on day 22. He was afebrile and did not require supplemental oxygen. Computed tomography of the chest revealed multiple bilateral lung nodules with bibasilar atelectasis and a ground glass opacity at the right lung base. There was no pulmonary embolism. An NP swab was again positive for SARS-CoV-2 by RT-PCR. He was admitted for intravenous fluid hydration and monitoring, but soon became persistently febrile and required 1-3 liters of supplemental oxygen. On day 29, repeat NP and sputum samples were both positive for SARS-CoV-2 by RT-PCR.

On day 30, serological testing did not detect antibodies to SARS-CoV-2 (EUROIMMUN anti-SARS-CoV-2 IgG ELISA), and remdesivir was initiated (10 day course). He received convalescent plasma therapy on day 31 and defervesced soon thereafter. His sputum remained positive for SARS-CoV-2 by RT-PCR on days 33 and 38. His condition continued to improve and he was discharged to home on day 39 of his illness.

The patient was without significant complaints at a follow-up telemedicine visit on day 60. Repeat serologic testing 66 days after his initial symptoms detected IgG antibodies to SARS-CoV-2 (Roche Elecsys Anti-SARS-CoV-2, outside hospital report). He also had three outpatient NP swabs at another institution on days 46, 57 and 66 that were all positive (Roche assay). While the patient remained positive for SARS-CoV-2 RNA on day 81 (Abbott assay), the decision was made to reinitiate lymphoma treatment given his relatively high Ct values, apparent seroconversion, and progression of his underlying disease. Chemotherapy was started on day 85 and completed on day 106. He had mild upper respiratory symptoms on day 106, and an NP swab was positive for SARS-CoV-2 by RT-PCR (Diasorin assay).

On day 119, patient presented to the emergency department with fevers, cough and shortness of breath. A chest radiograph showed new bilateral air space opacities. He was started on broad spectrum antibiotics. Two days after admission, he had a worsening chest radiograph and increasing oxygen requirement. On day 122, due to worsening symptoms and persistent fevers, the patient was given a second course of remdesivir (10 days) and convalescent plasma. He defervesced within 24 hours and was slowly weaned off of oxygen. He was discharged on day 131 of his illness.

### Laboratory Investigation

Residual respiratory tract specimens from days 7, 12, 22, 29 (one NP swab and one sputum), 33, 38, 81, 93, 106, and 119 (Figure 1) were recovered from the hospital clinical microbiology laboratory and cultured on Vero E6 cells. Cultures of all samples except day 81 produced cytopathic effect. Cells infected with supernatants from these primary cultures were positive for the SARS-CoV-2 nucleocapsid protein (Figure 1B), demonstrating the presence of infectious virus from days 7 through 119 of the patient’s illness.

We amplified and sequenced SARS-CoV-2 from RNA in the original NP and sputum specimens. A whole genome phylogenetic analysis with 100 SARS-CoV-2 sequences (Supplemental Table 1) from the same hospital revealed clustering of all nine sequences from this patient, which essentially rules out re-infection (Figure 2A). Six of the seven sequences from days 7-38 had an identical consensus. The NP sample from day 29 had two additional substitutions, one synonymous and one nonsynonymous (C5184U, ORF1a P1640L, Figure 2B). Given that the other day 29 sample was from sputum, these differences might reflect two different subpopulations present within the same host on that particular day. Four additional nonsynonymous substitutions were fixed by day 93. One of the nonsynonymous changes, ORF1a P1640F, actually had two mutations in the same codon –

**Figure 2:**
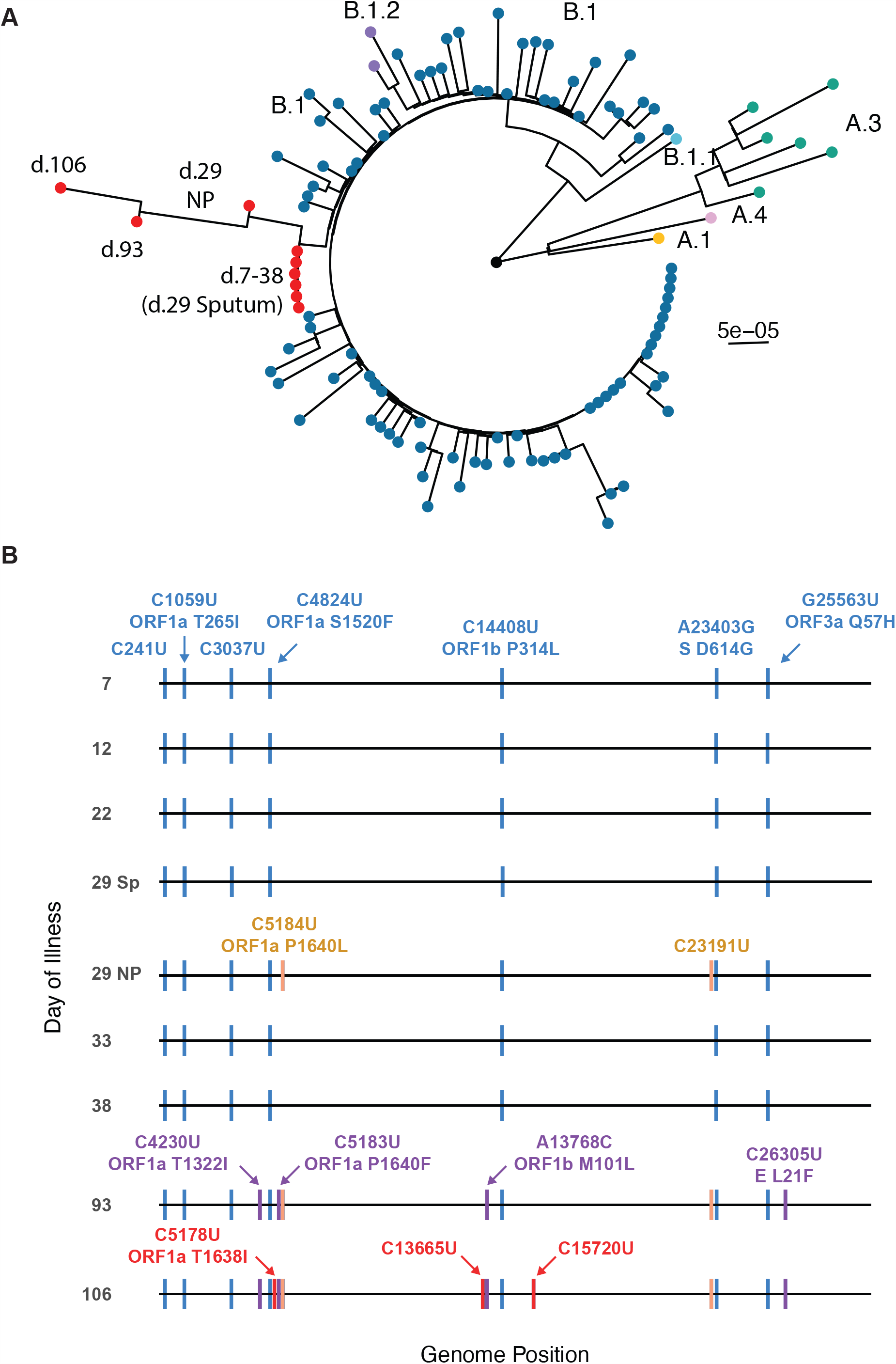
(A) Maximum likelihood phylogenetic tree of whole genome consensus sequences for nine samples from this case (red tips) and 100 other samples from inpatients at the same hospital (see Supplemental Table 1, color coded by lineage). Tree is rooted to Wuhan-Hu-1. Boostrap support (1000 replicates) at nodes for the samples from the case and for all lineages are > 90. (B) Consensus mutations in the nine samples sequenced. All nucleotide substitutions are shown. Amino acid substitutions are indicated for all nonsynonymous substitutions. Mutations in blue were in the original day 7 sample and persisted. Mutations in orange were detected in one of two day 29 samples and then at days 93 and 106. Mutations in purple were first detected in the day 93 sample and persisted at day 106. Mutations in red were fixed in the day 106 sample only. S = spike, E = envelope, ORF = open reading frame.

C5184U, which was fixed in the day 29 NP specimen, and C5183U, which was a minority variant in the day 29 and day 33 sputum samples (Supplemental Figure 1). The day 106 sample had one additional nonsynonymous substitution (C5178U, ORF1a T1368I), which was also present as a minority variant in the day 29 and 33 sputum samples. These data demonstrate the within-host evolution, and therefore replication, of SARS-CoV-2 over a four month period.

We evaluated the patient’s serological response with banked sera from days 30, 88, 120, and 122 (Supplemental Table 2). On day 30, he was negative for total antibodies against nucleocapsid (Roche), total antibodies against the spike receptor binding domain (Siemens), IgG against spike S1/S2 (Diasorin), and IgG against S1 (EUROIMMUN). He was categorically positive for nucleocapsid antibodies on day 60 (outside report) and only marginally positive on day 88 (index value of 2.1, threshold of 1.0 where values during natural infection are often >100); he remained negative for anti-spike antibodies although the Siemens index value after receiving plasma was numerically greater than baseline. It is not clear whether this represents seroconversion or decay of antibodies administered in the convalescent plasma. He was negative for all antibodies on day 120 and then again became marginally positive for nucleocapsid antibodies after the second administration of convalescent plasma.

## Discussion

Persistence of viral RNA and long-term replication are increasingly recognized as sequelae of acute viral infections ^7^. In most cases, these phenomena are attributable to incomplete immune clearance and/or ongoing viral replication in immune-privileged sites. For example, persistence of measles virus in the central nervous system has been well documented months after infection and Ebola virus replication has been found in the testes and retina ^8–10^. There are many examples of chronic infections with norovirus and influenza virus in individuals with adaptive immune deficits ^11,12^, and patients with common variable immune deficiency or agammaglobulinemia have been found to shed the oral poliovirus vaccine for many years ^13,14^.

Our patient’s mantle cell lymphoma, cancer chemotherapy, and associated immunosuppression are likely the reason for his protracted clinical course. While his regimen included broadly active, cytotoxic agents that would affect multiple aspects of immune function (e.g. doxorubicin and cyclophosphamide), we hypothesize that antibody-mediated ablation of B-cell precursors by mosunetuzumab and polatuzumab vedotin was primarily responsible for his prolonged viral shedding. The impact of both agents is expected to be greater on naïve – as opposed to memory – B-cells, which would be required for the initial response against a new pathogen, such as SARS-CoV-2. This humoral defect explains the absence of detectable seroconversion and recrudescence with reinitiation of his lymphoma treatment. The fact that he subsequently improved with each course of convalescent plasma (and remdesivir) demonstrates the important role that antibodies play in clearance of SARS-CoV-2.

Repeated sampling over a prolonged infectious period allowed us to examine the evolutionary dynamics of SARS-CoV-2 in a human host. Studies of noroviruses and influenza viruses in immunocompromised hosts suggests that these individuals may be reservoirs of antigenically novel viruses and that within-host selection may parallel evolutionary dynamics on larger scales ^15,16^. Prior to the first course of remdesivir and convalescent plasma (days 7-31), we observed a steady accumulation of minority single nucleotide variants (Supplemental Figure 1). Mutations accumulated over the next two months, and nine mutations fixed between days 93 and 106. None of these mutations have been observed at > 0.25% frequency in more than 88,000 sequences available in GISAID (Supplemental Table 3). Notably, there were no new mutations in the spike protein between days 7 and 106, despite early treatment with convalescent plasma. The ORF1b M101L substitution (day 93 and 106 samples) maps to NiRAN domain of the RNA dependent RNA polymerase, but is not known to mediate remdesivir resistance.

At this early stage of the pandemic, decisions regarding patient isolation and cohorting have largely been based on the results of nucleic acid testing. However, it is increasingly clear that SARS-CoV-2 RNA can be detected in clinical specimens long after a patient becomes culture negative, and likely non-infectious. This is consistent with studies of other respiratory viruses ^17 18^. In a comprehensive study of SARS-CoV-2 shedding in nine longitudinally sampled patients, W⍰lfel et al. were not able to culture virus 8 days after symptom onset, despite the persistence of viral RNA detectable by RT-PCR ^19^. This trend has since been found in multiple studies, and culture positivity has not been identified past 20 days ^4,20^. Because RT-PCR positivity appears to persist for several weeks, national guidelines have suggested that viral RNA detected after two weeks of illness is of unclear significance and favor basing isolation decisions on time since symptom onset (www.cdc.gov). Our case highlights the unresolved issue of immunocompromised hosts, who may shed infectious virus for longer periods of time and may require alternative criteria. While routine use of viral culture is currently not feasible, it may be reasonable to use Ct thresholds, RT-PCR for replicative subgenomic RNA, or seroconversion with titer as surrogates for the presence or absence of infectious virus.

Our case illustrates that some patients with defective immune responses can shed infectious virus for many more weeks than originally thought. This has important public health and infection control implications. A major limitation of our study is that it describes a single individual whose clinical course may not be broadly generalizable to other immunocompromised population. Nevertheless, we expect that additional cases such as the one described above will continue to elucidate important aspects of SARS-CoV-2 pathogenesis, evolution, and immunity.

## Data Availability

All data is available. Links and accession numbers are provided in the manuscript text file.

## Funding Acknowledgement

This work was supported by a COVID-19 Response Innovation Grant from the University of Michigan. Additional support for the enrollment of hospitalized patients with COVID-19 was provided by the CDC (U01 IP000974).

## Figure Legends

Supplemental Figure 1: Frequencies of seven single nucleotide variants (colored lines) that were identified in ≥ 2 samples: ORF1a C5178U, T1638I; ORF1a C5183U P1640S; ORF1a C6033U, A1923V; ORF1a C10702U, Synonymous; ORF3a G25644U, Synonymous; ORF8 G28239U, V116F, ORF8 C28253U, Syn. Pairwise nucleotide diversity (black line) in each sample is plotted by day of illness, second y-axis. Remdesivir was initiated on day 30 and convalescent plasma was given on day 31.

Supplemental Table 1: GISAID accession numbers for sequences in Figure 2

Supplemental Table 2: Serological testing

Supplemental Table 3: Frequency of case patient viral mutations in GISAID database

## Supplementary Methods

All experiments using SARS-CoV-2 were performed at the University of Michigan under Biosafety Level 3 (BSL3) protocols in compliance with containment procedures in laboratories approved for use by the University of Michigan Institutional Biosafety Committee (IBC) and Environment, Health & Safety (EHS). This study and the use of residual specimens from patients with COVID-19 have been approved by the University of Michigan Institutional Review Board (IRB).

All RT-PCR testing and associated cycle threshold values were based on assays performed in the Michigan Medicine Clinical Microbiology Laboratory. “Diasorin” refers to the Simplexa COVID-19 Direct assay, level of detection 500 copies per ml. “Abbott” refers to the Real Time SARS-CoV-2 assay, level of detection 100 copies per ml.

Vero E6 cells were purchased from ATCC® (CRL-1586™) and maintained at 37°C with 5% CO2 in Dulbecco’s Modified Eagle’s Medium (DMEM, Gibco) supplemented with 10% heat-inactivated fetal bovine serum (FBS), HEPES, non-essential amino-acids, L-glutamine, and 1x Antibiotic-Antimycotic solution (Gibco).

Nasopharyngeal swab and sputum specimens were transferred to a BSL3 laboratory and centrifuged at 1200g for 10 minutes to remove cellular debris and contaminants. Two hundred microliters of sample were added to Vero E6, allowed to adhere overnight on a 12-well plate and maintained in DMEM supplemented with 2% FBS. Cultures were checked daily for cytopathic effect by microscopic inspection. Cells and supernatants were harvested and kept at −80°C. The SARS-CoV-2 WA1 strain was obtained from BEI Resources and was propagated in Vero E6 cells.

For immunostaining, 384-well plates (PerkinElmer, 6057300) were seeded with Huh-7 cells at 3000 cells/well and allowed to adhere overnight. The plates were then transferred to BSL3 containment where supernatants of clinical isolate-infected Vero E6 cells were added to the cells (final concentration of 10%) and incubated at 37°C for 48 h. Huh-7 cells infected with SARS-CoV-2 WA1 at a multiplicity of infection (MOI) of 0.2 were used as positive control of infection. Two days after infection, cells were fixed with 4% PFA for 30 minutes at room temperature, permeabilized with 0.3% Triton X-100, and blocked with antibody buffer (1.5% BSA, 1% goat serum, and 0.0025% Tween 20). The plates were then sealed, surface decontaminated, and transferred to BSL2 for staining. To detect viral-infected cells, anti-nucleocapsid protein (anti-N) SARS-CoV-2 antibody (Antibodies Online, Cat# ABIN6952432) was used as a primary antibody with an overnight staining at 4°C followed by staining with secondary antibody Alexa-647 (goat anti-mouse, Thermo Fisher, A21235). Hoechst-33342 pentahydrate (bis-benzimide) was used for nuclei staining (Thermo FIsher, H1398). Plates were then imaged with a Thermo Fisher CX5 high content microscope with a 20X/0.45NA LUCPlan FLN objective with LED excitation (386/23nm, 485/20nm, 560/25nm, 650/13nm).

Viral genomic RNA was extracted from the original nasopharyngeal swab and sputum specimens using Direct-Zol RNA miniprep kits (Zymo Research). Complementary DNA corresponding to the genome was amplified by RT-PCR in two multiplex reactions using the ARTIC network V3 primer set. Sequencing libraries were prepared using the NEBNext Ultra II kit and sequenced on an Illumina MiSeq with 2×250bp reads, V2 chemistry. Reads were aligned to the Wuhan-1 reference (GenBank: MN908947.3) with BWA-MEM version 0.7.15. Sequencing adaptors were removed and the ARTIC primer were trimmed with iVar version 1.2.1. We determined the consensus sequences with samtools (version 1.5) mpileup and iVar consensus, excluding bases with a quality score below 20. Positions with fewer than 10 reads were given an N. Insertions and deletions were manually curated by inspecting sequence reads in IGV (version 2.8.0). Single nucleotide variants were identified using iVar 1.2.1 with a minimum sequencing depth of 400 and a frequency threshold of 2%. Variant bases with quality score less than 30 and mapping quality score less than 20 were excluded. We accounted for strand bias by performing a two-sided Fisher’s exact test that tests the hypothesis that the forward/reverse strand counts in the minor base are derived from the same distribution as the consensus base at that position. A Bonferroni multiple test correction was applied and variants with an adjusted p-value < 0.05 were excluded.

Whole genome consensus sequences were aligned to the Wuhan-Hu-1 reference with MUSCLE (version 3.8.31) and the phylogeny constructed with IQ-TREE using a GTR model and 1000 ultrafast bootstrap replicates. To determine the frequency of case patient mutations in the larger global dataset of SARS-CoV-2, we downloaded all sequences >27000 bases in length from GISAID and aligned them with MAFFT within the nextstrain environment (Supplemental Table 3). Sites were queried individually and sequences with a gap or N at a given site were excluded.

Seroassays were performed on banked serum using the indicated kits according the manufacturers’ protocols.

All analysis code, patient sequences, and metadata are available https://github.com/lauringlab/ProlongedReplicationCase. Original sequence reads are available at www.ncbi.nlm.nih.gov/sra under Bioproject PRJNA662589 (release date 9/23/2020).

## Notes

### Competing Interest Statement

The authors have declared no competing interest.

### Author Declarations

Determined exempt by University of Michigan IRB. Residual clinical specimens were obtained under a protocol approved by the University of Michigan IRB.

